# Acceptability and preliminary effectiveness of a remote dementia educational training among healthcare professionals

**DOI:** 10.1101/2022.01.25.22269850

**Authors:** Jaime Perales-Puchalt, Ryan Townley, Michelle Niedens, Eric D Vidoni, K Allen Greiner, Tahira Zufer, Tiffany Schwasinger-Schmidt, Jerrihlyn L McGee, Hector Arreaza, Jeffrey M Burns

## Abstract

**Background:** Optimal care for families living with Alzheimer’s disease and related disorders (ADRD) has the potential to improve their lives. However, ADRD care remains under-implemented among healthcare professionals, partly due to professionals’ limited ADRD training and inexperience. Professional training might help, but most training is in person, time-intensive, and does not focus on the potential of early detection, client empowerment, and cultural competency. We aimed to explore the acceptability and preliminary effectiveness of an online ADRD training, The Dementia Update Course, which addressed these issues. We hypothesized that the Dementia Update Course would lead to increased levels of perceived ADRD care competency among healthcare professionals.

**Methods:** This was a mixed-methods research design using pre-post training assessments. The training included 59 primary care providers (PCPs) and other healthcare professionals (e.g., medical specialists, nurses, social workers). The Dementia Update Course was a remote 6.5-hour training that included didactic lectures, case discussion techniques, and materials on ADRD detection and care. Outcomes included two 5-point Likert scales on acceptability, eleven on perceived dementia care competency, and the three subscales of the General Practitioners Confidence and Attitude Scale for Dementia. We used paired samples t-tests to assess the mean differences in all preliminary effectiveness outcomes.

**Results:** The training included 18.0% of professionals that self-identified as non-White or Latino and 37.7% of professionals who served in rural areas. Most participants (90.0% and 87.5%) reported a high likelihood to recommend the training and high satisfaction respectively. All preliminary effectiveness outcomes analyzed in the total sample experienced a statistically significant improvement from pre- to post-training averaging 0.7 points in 1-5 scales (p<0.05). Most outcomes improved statistically among PCPs too.

**Conclusions:** A relatively brief, remote, and inclusive ADRD training led to high levels of acceptability and improved perceived ADRD care competency among PCPs and other healthcare professionals. Future research should include a control group and assess guideline compliance, behavioral outcomes, and health outcomes among people with ADRD and their families.

## Introduction

Alzheimer’s disease and related disorders (ADRD) pose a serious public health threat worldwide. The population aged 65 and older is increasing and the risk of ADRD is known to increase with age (Administration on Aging et al., 2016; Hebert et al., 2013). According to the Pan American Health Organization, ADRD was the second leading cause of mortality among people 18 and older in the US in 2019, and the 15^th^ leading cause of disability (Pan American Health Organization, 2021). Individuals with ADRD have more chronic conditions (e.g., diabetes, depression, pneumonia), polypharmacy use, and hospitalizations than older adults without ADRD (Alzheimer’s Association, 2021). Caregivers of people with ADRD often experience disproportionate depression (∼34%) and anxiety (∼44%) vs caregivers of people with stroke (19% of depression and 31% of anxiety) and non-caregivers of similar ages (∼12% of depression) (Alzheimer’s Association, 2021; Sallim et al., 2015). ADRD costs exceed those of cancer and heart disease by $32 and $7 billion respectively (Alzheimer’s Association, 2021; Hurd et al., 2013).

Early diagnosis and implementation of optimal care can improve the prognosis for people with ADRD and is a priority for the National Alzheimer’s Project Act (NAPA) (U.S. Department of Health & Human Services, 2016). Though there are no treatments that prevent or stop the progression of ADRD, there is evidence that pharmacologic and non-pharmacologic interventions can stabilize and delay the progression of cognitive, functional, and behavioral outcomes, improving the lives of individuals with ADRD and their families (Cotter, 2006; Gitlin et al., 2012). Given the progressive nature of ADRD, early diagnosis may allow both the person with ADRD and the family to participate in their care plan and begin more efficacious interventions at an earlier time point (Milne et al., 2008). Early detection and care have benefits at the individual, familial and societal levels (Black et al., 2018; Mesterton et al., 2010; Wimo, 2004; Winblad & Wimo, 1999). For example, longitudinal data from the Medicare fee-for-service claims suggests that individuals with ADRD who were newly diagnosed and treated had a lower mortality, institutionalization rate, and annual costs than those who were not treated (Black et al., 2018). The healthcare system is ideally positioned to coordinate ADRD care, as most older adults in the US are insured and have a usual source of healthcare (Administration for Community Living, 2015; U.S. Department of Health and Human Services, 2017). Among healthcare professionals, primary care providers (PCPs) have the potential to play an important role as they are often the first point of contact (Drabo et al., 2019).

Despite the potential of healthcare professionals to improve the lives of families with ADRD, care remains under-implemented. Approximately 49.2%-56.5% of Americans with ADRD are unaware of their condition and remain potentially undiagnosed (Lin et al., 2020). In the clinic, only 16% of people 65 and older receive regular cognitive assessments during routine health check-ups (Alzheimer’s Association, 2019). Only about 51% of healthcare providers follow up with cognitive screening results reported by their patients after a screening event (Galvin et al., 2020), half of PCPs recommend laboratory testing for all patients with a detected cognitive impairment, and 17% percent make specialist referrals for all patients with a detected cognitive impairment (Alzheimer’s Association, 2019). Half of people with ADRD are being treated with cognitive medications (Koller et al., 2016). Almost 80% of family caregivers of people with ADRD report unmet needs in at least one service area (e.g., activities of daily living, ADRD symptoms, timing of care), and nearly one third do not receive any type of caregiver support services (Li, 2012; Scharlach et al., 2008).

In addition, Latino, non-Latino Black, and rural individuals have a higher ADRD risk and are disproportionately underserved in care outcomes (Alzheimer’s Association, 2019; Bédard et al., 2004; Koller et al., 2016; Lin et al., 2020; Scharlach et al., 2008; Weden et al., 2018). For example, the likelihood of non-Latino Black and Latino individuals with ADRD to be undiagnosed is 34-40% higher than non-Latino Whites (Lin et al., 2020). If diagnosed, Latinos with ADRD experience a delay in their diagnosis of eight months compared to non-Latino Whites (Barker et al., 2005). Non-Latino Black people with ADRD are 6% less likely to use anti-dementia medications than non-Latino Whites (Koller et al., 2016). Non-Latino White caregivers have been found to be 2.3 times more likely than foreign-born Latinos to use ADRD support services (Scharlach et al., 2008). People with ADRD residing in rural settings are less likely to receive formal supports than those in urban settings (35% vs 88% respectively) (Bédard et al., 2004).

Multiple factors contribute to the under-implementation of ADRD detection and care in healthcare. Strategies to provide early ADRD diagnosis and care in primary care clinics are lacking (Chodosh et al., 2007; Rosen et al., 2002). Some healthcare providers are reluctant to address ADRD due to limited ADRD knowledge on the potential benefits of appropriate diagnosis and care (Boise et al., 1999; Bradford et al., 2009). Some providers also lack the necessary tools and resources to diagnose, provide care for or refer to frequently fragmented community resources (Boise et al., 1999; Bradford et al., 2009). These barriers are more prevalent among non-Whites and Latinos, as many providers lack cultural and linguistic proficiency and knowledge of validated tools for assessment and care (Boise et al., 1999; Ferraro, 2015; Ganguli et al., 2018; Ortiz & Fitten, 2000; Vega et al., 1999).

Training healthcare professionals can address some of the barriers to ADRD care implementation. Few ADRD training programs for healthcare professionals exist and most have been developed and tested in the last decade (Costa et al., 2019; Perry et al., 2011). However, most training programs have relied on in-person sessions, which pose barriers to largely rural regions such as the Midwestern US. This barrier and the COVID-19 pandemic have also highlighted the importance of virtual or remote training programs. Most existing training programs are time-intensive and add to the already burdened schedules of healthcare professionals (Boise et al., 1999; Bradford et al., 2009). Few training programs focus on early detection and empowering the person with ADRD to participate in the decision-making process of their care, which potentially reduces the probability of beneficial health and economic outcomes (Mesterton et al., 2010; Polese et al., 2016; Wimo, 2004; Winblad & Wimo, 1999). The focus on health disparities within most ADRD healthcare professional training programs is scarce, which may contribute to widening these disparities. There is a need for ADRD training programs for healthcare professionals that are highly accessible, time-efficient, include early detection, center the care on the patient and addresses ADRD disparities. The objective of the current manuscript was to explore the acceptability and preliminary effectiveness of a relatively brief online ADRD training that addresses these needs, the Dementia Update Course. We hypothesized that the Dementia Update Course would lead to increased levels of ADRD care competency among PCPs and other healthcare professionals (e.g., medical specialists, nurses, social workers) in general and specifically competency to serve non-White and Latino people with ADRD and their families.

## Methods

This study used a mixed-methods concurrent design. The quantitative method dominates, while the qualitative method is nested within. The quantitative method included pre-post-training assessments, whereas the qualitative method included a qualitative descriptive approach. This healthcare professional training was part of two NIH-funded projects (R24 AG063724 & K01 MD014177). First, MyAlliance is a three-year project that aims to increase ADRD research participation using a three-pronged strategy. This strategy involves the engagement of healthcare professionals, community organizations, and the lay community. MyAlliance has a special interest in increasing ADRD research participation among rural, non-White, and Latino communities. Second, Alianza Latina (Latino Alliance) is a five-year project that aims to improve ADRD care services among Latinos. The Alianza Latina project aims to improve healthcare professionals’ ability to detect Latinos with ADRD and provide more appropriate care and referrals to a culturally and linguistically proficient ADRD care manager. The University of Kansas Medical Center Institutional Review Board deemed this project a quality improvement project and did not require that participants complete an informed consent.

The Dementia Update Course was a one half-day (6.5 hours) educational event held three times throughout 2021 (March, July, and September). The course was open to a wide diversity of professionals, even though only physicians, nurses, and social workers could apply for continuing education credits upon attendance to 90% of the sessions. The cost of the training was $50, unless healthcare professionals requested a waiver, in which case, there was no cost. The educational framework that informed the Dementia Update Course was a combination of the Health Belief Model and Social Learning Theory, by including clinically-focused lectures, case examples with discussion, videos and materials for additional reference (Rosenstock et al., 1988). Table 1 shows the agenda, which includes ADRD detection and diagnosis, treatment, and care for cognitive and behavioral symptoms, and how to apply the tools to a healthcare professional’s daily workflow. Modules highlighted the importance of early detection, patient empowerment, cultural competence, and tailoring care to disparity populations, especially Latinos. Elements of patient empowerment followed the Client Empowerment Model, which is in concert with strength-based social work philosophy, and incorporates components of knowledge/health literacy, shared decision making, personal control, and positive patient-professional interaction based on the stage of ADRD (Polese et al., 2016). Cultural competence included information about disparities in ADRD experienced by diverse groups, and how they can be addressed in the healthcare practice. Information integrated key elements of culturally and linguistically appropriate services (CLAS), covered patient-centered care and effective communication, and adapted and validated tools with a special focus on Latinos (Martinez et al., 2004; US Department of Health & Human Services, 2018). The training was conducted live and online via videoconferences. All attendees also received copies of recommended cognitive screening tools and the materials reviewed during the course. Staff from the University of Kansas Alzheimer’s Disease Research Center (KU ADRC) led all the modules. Members of the KU ADRC recruited participants via emails to clinics, phone calls, or word of mouth.

**Table 1.** The Dementia Update Course agenda.

|  |  |
| --- | --- |
| 8:00 – 8:30 a.m. | Brief overview to KU Alzheimer's Disease Center |
| 8:30 – 8:50 a.m. | Shifting to a new model of dementia care |
| 8:50 – 9:20 a.m. | Recognizing early clues, screening tools |
| 9:20 – 9:30 a.m. | Stretch Break |
| 9:30 – 10:20 a.m. | Dementia Work- Up: When, What, Why and Differential Diagnosis |
| 10:20 – 10:30 a.m. | Stretch Break |
| 10:30 – 11:00 a.m. | Latinos and Dementia |
| 11:00 – 12:00 p.m. | Treating the Person with a Dementia: Disclosure, Early Intervention, and Medication |
| 12:00 – 1:00 p.m. | Lunch Break |
| 1:00 – 2:00 p.m. | Behavioral and Affective Challenges in Dementia |
| 2:00 – 2:30 p.m. | Driving |
| 2:30 – 2:40 p.m. | Stretch Break |
| 2:40 – 3:40 p.m. | Latest Alzheimer's Research and integration of research into your practice |
| 3:40–4:10p.m. | Dementia Capable Workflow and Environmental Considerations |
| 4:10 – 4:30 p.m. | Final Questions/Post Test |

### Procedure

A staff member of the KU ADRC sent an email to participants one day before the training asking them to complete a pre-training survey via REDCap (Harris et al., 2009), and highlighting the voluntary and anonymous nature of the assessment. On the day of the training, one of the instructors introduced the training content to the audience and the instructors presented their modules, requesting recurrent interaction. Immediately after finishing the training, a staff member reminded the participants to complete the post-survey, which a staff had emailed them one hour before the end of the training.

### Data Collection and Measures

Pre-training survey socio-demographic information included age, gender, race, ethnicity, occupation, rural/urban setting of their clinic, and state. Post-training acceptability of the intervention was measured by asking two items developed by the research team that included 5-point Likert scales about how likely they are to recommend the training to a colleague, and how satisfied they were with the training from 1 (not at all) to 5 (very much). The post-training survey also included an open-ended acceptability question asking what participants liked most about the training.

Preliminary effectiveness outcomes included several ADRD perceived competency constructs and other ADRD-related attitudes measured via the following survey instruments. The first was the General Practitioners Confidence and Attitude Scale for Dementia (GPACS-D) comprises three subscales: Confidence in Clinical Abilities (six items), Attitude to Care (six items), and Disengagement (three items). The scale has been validated using confirmatory factor analysis, reliability, validity, and sensitivity to change tests (Mason et al., 2019; Mason et al., 2020). Each item response ranges from 1 (strongly agree) to 5 (strongly disagree). We divided each subscale score by the number of their number of items to standardize the score ranges from 1 to 5. Higher scores mean higher confidence in their clinical abilities, better attitudes towards ADRD care, and higher disengagement with ADRD care. We adapted the GPACS-D by using US English spelling and replacing the community organization “Alzheimer’s Australia” with the “Alzheimer’s Association”. In our study, the Cronbach Internal Consistency Alpha was 0.8 for both Confidence in Clinical Abilities and Attitude to Care, and 0.7 for Disengagement.

The second instrument included eleven items developed by the research team. These items included 5-point Likert scales about perceived confidence in explaining the importance of early identification of cognitive decline, selecting the appropriate tools to screen for early signs of ADRD, differentially diagnosing the most common signs of ADRD, identifying the appropriate interventions based upon identification of cognitive decline, using elements of the Client Empowerment Model as applied to ADRD, implementing cognitive screenings in regular workflow, impacting the quality of life of families with ADRD, and serving English-speaking Latino, Spanish-speaking Latino, Non-Latino Black and other non-White families with ADRD. Each item response ranged from 1 (not at all) to 5 (totally confident).

### Analysis

Participants from diverse professional backgrounds were eligible to participate in the course. However, for the analysis, only those with a healthcare professional background were eligible. This includes physicians, physician assistants, and nurse practitioners, other nurses, nursing assistants, behavioral providers, personal care attendants, information and referral providers, options counselors, social workers, case managers, and related professions. Analyses were conducted for the total sample and stratified into PCPs and other healthcare professionals. The reason for this stratification was that PCPs have the potential to play an important role in ADRD care, as they are often the first point of contact (Drabo et al., 2019). Means, standard deviations, percentages, and frequencies were calculated for pre-training survey characteristics and acceptability outcomes. Differences in baseline characteristics between participants who completed the post-training survey and those who did not were calculated using Chi-Square tests and independent samples t-tests. Paired samples t-tests were used to compare the healthcare professionals’ perceived confidence in serving Spanish-speaking Latino vs English-speaking Latino and non-Latino Black families with ADRD. Paired samples t-tests were conducted to assess the mean differences in all preliminary effectiveness outcomes. We only conducted analyses of diagnosis-related outcomes (GPACS-D and three confidence items) among PCPs, given that the other professional groups are diverse, and most do not diagnose ADRD. We used SPSS 20 to run quantitative analyses and used a significance level of α=0.05 to protect against type I error. To analyze the answers to the open-ended acceptability question, we coded qualitative answers and later defined acceptability categories that summarized different discourses.

## Results

### Baseline characteristics of the sample

The characteristics of the sample are shown in Table 1. Seventy-three individuals RSVP’d and 59 individuals attended one of the three training sessions and responded to the pre-training survey. A total of 53 participants were eligible for analysis, and 41 (77.4%) completed the post-training survey. Those excluded reported no profession, or were recruitment professionals, research assistants or research interns. Nearly all participants were from Kansas or Missouri. The average age of participants was 48.2 (SD 14.6) and 43 participants were women (81.1%). Most participants were non-Latino White (n=41; 82.0%) although Latinos, non-Latino Blacks, Non-Latino Asians, and Non-Latino Mixed-race individuals were also represented. Twenty participants (37.7%) reported they practiced in a rural community.

Participants’ professions included PCPs (n=17; 32.1%), physicians, nurses, and related professions not in primary care (n=15; 28.3%), case managers, care coordinators, and discharge planners (n=12, 22.6%), social workers, and social work interns (n=5, 9.4%), and information, and referral specialists (n=4, 7.5%). Participants who completed the post-training survey were more likely to be older than those who did not (mean age= 50.4 vs 40.6; p=0.038). Participants’ perception of their ability to serve Spanish-speaking Latino families with ADRD (2.0; SD: 0.9) was lower than that of English-speaking Latino (2.9, SD: 1.0; p<0.001) and non-Latino Black families (3.0; SD: 1.1; p<0.001). There was no difference between their perceived ability to serve English-speaking Latino and non-Latino Black families with ADRD (p=0.109).

**Table 2.** Sociodemographic characteristics of the healthcare professional sample.

| **Month of ADRD training in 2021** | **Total**<br>**(n=53)** | **Completers**<br>**(n=41)** | **Non-completers**<br>**(n=12)** | **P Value** |
| --- | --- | --- | --- | --- |
| **Date of training** |  |  |  | 0.104 |
| **March, % (n)** | 32.1% (17) | 39.0% (16) | 8.3% (1) |  |
| **July, % (n)** | 30.2% (16) | 29.3% (12) | 33.3% (4) |  |
| **September, % (n)** | 37.7% (20) | 31.8% (13) | 58.4% (7) |  |
| **Age in years, mean (SD)** | 48.2 (14.6) | 50.4 (15.7) | 40.6 (11.4) | 0.038 |
| **Women, % (n)** | 81.1% (43) | 82.9% (34) | 75.0% (9) | 0.403 |
| **Ethnoracial group** |  |  |  | 0.842 |
| **Non-Latino White, % (n)** | 82.0% (41) | 80.0% (32) | 90.0% (9) |  |
| **Latino (of any race), % (n)** | 8.0% (4) | 7.5 % (3) | 10.0% (1) |  |
| **Non-Latino Black, % (n)** | 4.0% (2) | 5.0% (2) | 0.0% (0) |  |
| **Non-Latino Asian, % (n)** | 4.0% (2) | 5.0% (2) | 0.0% (0) |  |
| **Non-Latino Mixed race, % (n)** | 2.0% (1) | 2.5% (1) | 0.0% (0) |  |
| **Practice in rural communities, % (n)** | 37.7% (20) | 36.6% (15) | 41.7% (5) | 0.501 |
| **Main profession** |  |  |  | 0.170 |
| **Primary care providers, % (n)** | 32.1% (17) | 39.0% (16) | 8.3% (1) |  |
| **Physicians, nurses, and related professions not**<br>**in primary care, % (n)** | 28.3% (15) | 22.0% (9) | 50.0% (6) |  |
| **Case managers, care coordinators and discharge**<br>**planners, % (n)** | 22.6% (12) | 24.4% (10) | 16.7% (2) |  |
| **Social workers and social work interns, % (n)** | 9.4% (5) | 7.3% (3) | 16.7% (2) |  |
| **Information and referral specialists, % (n)** | 7.5% (4) | 7.3% (3) | 8.3% (1) |  |

**Table 2.**
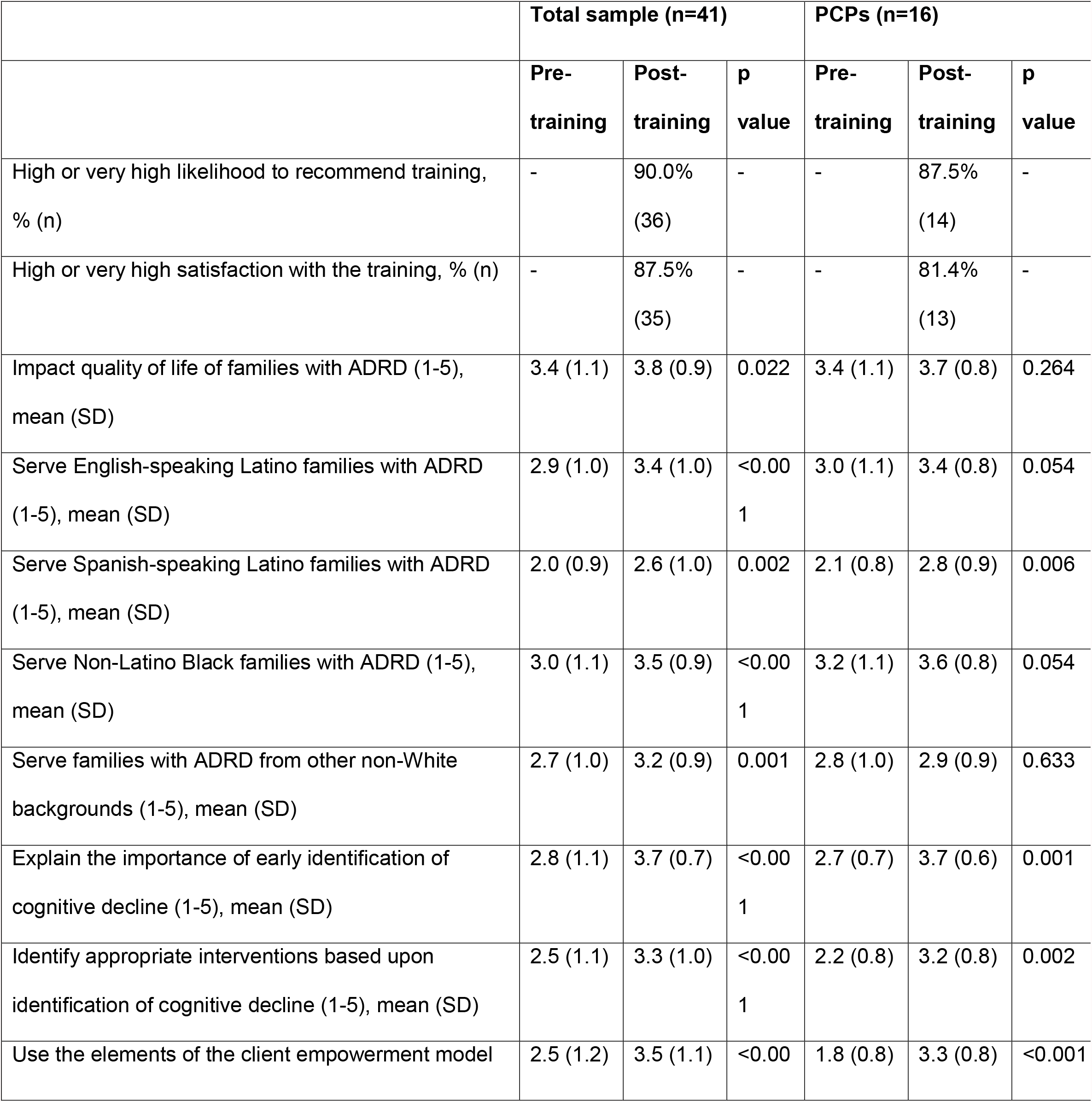
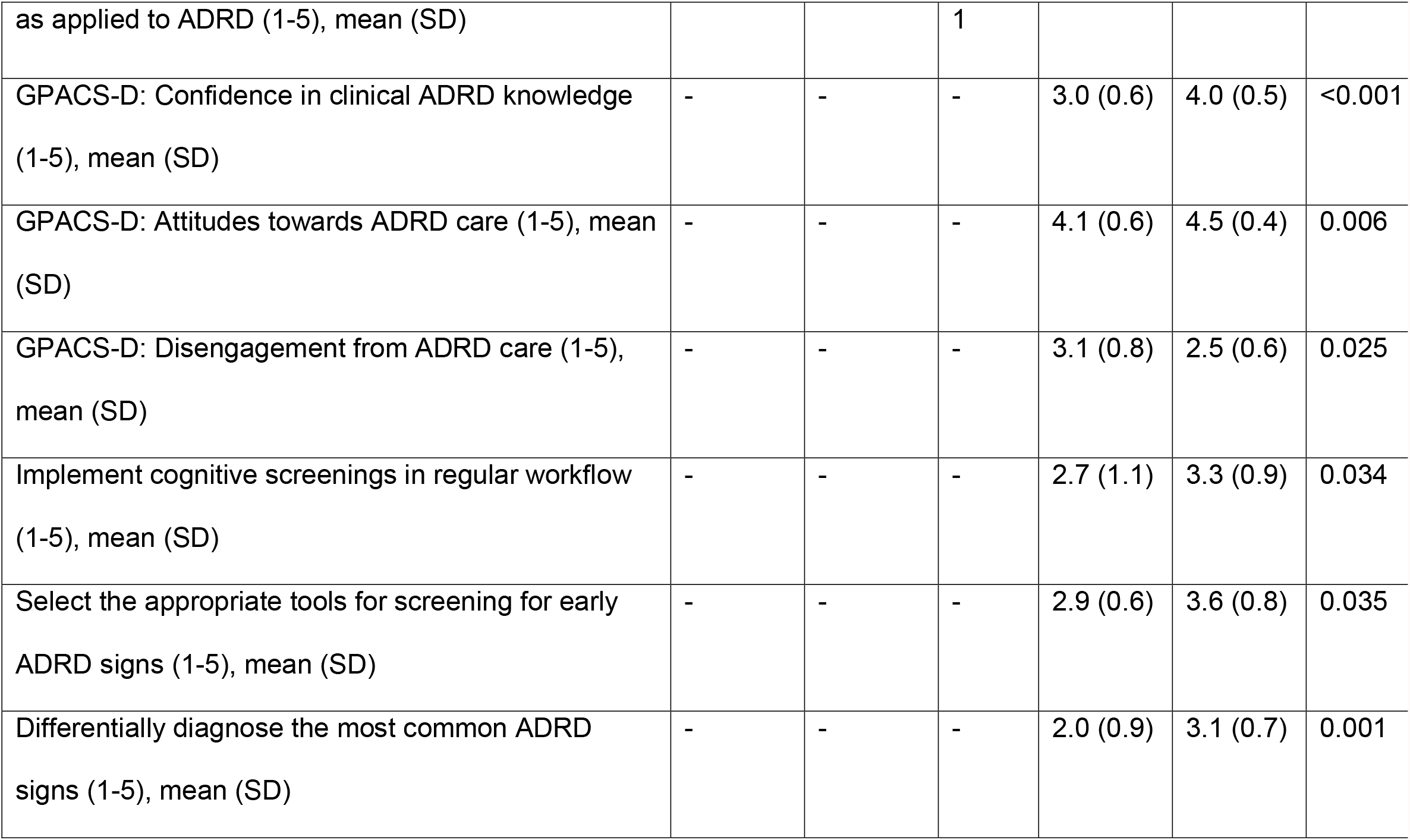
Acceptability and preliminary effectiveness of the ADRD training on healthcare professional outcomes.

### Acceptability and preliminary effectiveness of the Dementia Update Course

Table 3 shows the acceptability and preliminary effectiveness of the Dementia Update Course in the total sample and among PCPs. In the total sample, 36 participants (90.0% of responders) said their likelihood of recommending the training to a colleague was either high or very high, and 35 (87.5%) said their satisfaction with the training was high or very high. No participants rated their likelihood of recommending the training or their satisfaction as low or very low. All preliminary effectiveness outcomes experienced a statistically significant improvement from pre-training to post-training (p<0.05). These include improvements in their confidence to improve the quality of life of families with ADRD, explain the importance of early detection, identify appropriate interventions of cognitive decline, use the elements of the Client Empowerment Model, and serve families with ADRD of diverse ethnic and racial backgrounds. Preliminary effectiveness findings were similar among PCPs, although four out of the nine were not statistically significant. All preliminary effectiveness outcomes specific to PCPs (those that included diagnosis) experienced a statistically significant improvement from pre-training to post-training (p<0.05). These include improvements in their GPACS-D scores of their confidence in their clinical ADRD knowledge, attitudes towards ADRD care and disengagement from ADRD care, as well as their perceived confidence in implementing cognitive screenings in their regular workflow, select the appropriate tools for screening for early signs of ADRD, and differentially diagnose the most common signs of ADRD.

At the end of the training, participants responded to open-ended questions about what training aspects they perceived to be most useful. We categorized responses into four themes. First, participants showed gratitude and spoke of the usefulness of the training. An example of this category includes “Thank you for making a difference in the lives of those with dementia and their families!”. Second, participants highlighted the usefulness of training on current research and up-to-date care: “Learning about the KU Alzheimer’s Disease Center and the current knowledge and advancements regarding early detection” or “I found the ongoing research and explanations behind the diagnosis very helpful! Thank you!!”. Third, participants found different aspects of the teaching modalities used helpful: “Case ‘studies’, real-life examples” or “Driving discussion was the most helpful lecture. The specific example of what was needed and having an occupation therapist provide the exam was a helpful idea”. Fourth, participants highlighted the importance of topics including detection (imaging, screening tools, early detection), care (pharmacological and non-pharmacological), ADRD types and stages, and resources available.

## Discussion

The current manuscript aimed to explore the acceptability and preliminary effectiveness of the Dementia Update Course among PCPs and other professionals. In line with our hypothesis, findings suggest that this real-world training leads to improved ADRD-related attitudes, and perceived ADRD care competency among PCPs and other healthcare professionals. Previous research shows that training improves ADRD knowledge, attitudes, and perceived competence to different extents among healthcare professionals in the US and elsewhere (Bentley et al., 2019; Chodosh et al., 2006; Edwards et al., 2015; Galvin et al., 2012; Lathren et al., 2013; Lee et al., 2013; Mason et al., 2020; Vollmar et al., 2007; Wang et al., 2017). However, this training is innovative, as it focuses on eliminating health disparities, promoting early detection, and empowering the person with ADRD, which may bring about health and economic benefits to families with ADRD irrespective of their ethnic and racial background (Mesterton et al., 2010; Polese et al., 2016; Wimo, 2004; Winblad & Wimo, 1999). Moreover, the logistics of this training program address common and emergent barriers to ADRD training, including professionals’ burdened schedules, distance to training venues, and the COVID-19 pandemic.

A key component of the NAPA is to address ADRD care disparities among ethnically and racially minoritized populations (U.S. Department of Health & Human Services, 2016). To address health disparities, the Institute of Medicine recommends cultural competence training programs given their success in improving skills, knowledge and attitudes among healthcare professionals (Institute of Medicine, 2001). To our knowledge, this is among the first studies to include a cultural competence in an ADRD training curriculum. The Dementia Update Course led to similar increases in the perceived ability to serve families with ADRD of different ethnic and racial backgrounds. These similar improvements took place despite the fact that healthcare professionals were less confident about serving Spanish speaking Latinos, likely due to language, immigration status, and insurance status barriers (Administration for Community Living, 2015). Another important aspect is the representation of professionals from non-White and Latino backgrounds in our training sessions, who are under-represented in their professions, and can reduce health disparities when matched with patients of similar backgrounds (Cooper-Patrick et al., 1999; Cooper et al., 2003; King et al., 2004; Persky et al., 2013; Saha & Beach, 2020; Saha et al., 1999; Traylor et al., 2010). In our sample, nearly 20% of participants identified as a member of a group other than non-Latino White, which is a higher proportion than the physician workforce in Kansas (14.4%) (Association of American Medical Colleges, 2017).

Our inclusion of more than one-third of professionals who practiced in rural communities was encouraged by our region’s largely rural population and NAPA’s urge to facilitate the translation of research findings into rural public health practice (U.S. Department of Health & Human Services, 2016). In rural areas, ADRD specialists might not be readily available, and PCPs might be the only professionals in charge of ADRD detection and care (Galvin et al., 2012). To our knowledge, only one other ADRD training study has focused on rural healthcare professionals (Galvin et al., 2012). This training increased professionals’ knowledge and confidence to use cognitive screening tools to help diagnose and treat patients with ADRD. Our work builds upon this research by making it fully remote and reducing the number of days of training from three to one.

The current study has certain limitations. Our small non-probabilistic sample may limit the external validity of these findings. The sample only included individuals who decided to attend, potentially introducing selection bias leading to a sample with positive baseline ADRD care attitudes or receptiveness to the training. The study lacked a control group and randomization, thus preventing causality inference. It remains unknown whether changes in attitudes will translate into positive ADRD care behaviors. Previous studies suggest that it is possible to increase compliance with ADRD guidelines via similar training programs (e.g., increased use of cognitive screening tools) (Cherry et al., 2004; Lathren et al., 2013; Lee et al., 2013). Long-term professional outcomes of the Dementia Update Course will be reported in a future manuscript.

This study has significant implications for public health. The fact that this real-world training leads to improved perceived ADRD-related attitudes and perceived care competence indicates that relatively brief, remote training programs can have similar effects to more time-intensive in-person versions. The improvement of most outcomes specifically among PCPs is especially relevant. PCPs are the first point of contact with families with ADRD, and are in an optimal position to detect and initiate their care coordination (Drabo et al., 2019).

Studies should compare the cost-effectiveness and implementation outcomes of training programs such as the Dementia Update Course vs longer, more time-intensive, and in-person training programs. There was post-training survey attrition among physicians, nurses, and related professions not in primary care, but we did not assess reasons for such attrition. Future studies should explore these reasons to increase the feasibility of ADRD training programs among diverse healthcare professions. Future studies should expand understanding of ADRD specific curricula in primary care residencies and other programs through larger samples and a higher diversity of regions. Ultimately, studies may also want to assess the impact of these training programs on healthcare behavioral outcomes (e.g., compliance with guidelines), and directly on families with ADRD. This impact could be measured as effectiveness or cost-effectiveness by including comparison groups exposed to different types of training features (e.g., duration, in-person modality, topics).

## Conclusion

The Dementia Update Course, a relatively brief and remote ADRD training led to improved ADRD attitudes and perceived care competency among PCPs and other healthcare professionals. While other training programs have achieved similar outcomes, this training program addresses common barriers to ADRD care implementation by making it highly accessible, and including issues that are public health priorities, such as early detection, client empowerment, and addressing ADRD disparities via cultural competency (U.S. Department of Health & Human Services, 2016). Its high acceptability together with its relative brevity and virtual accessibility, make this training an ideal candidate for broad implementation.

## Data Availability

All data produced in the present study are available upon reasonable request to the authors

## Acknowledgements

The authors also thank Mary Beth Warren for assisting with the continuing education logistics, those who participated in the training program, and those who promoted it. The authors also thank everybody who has contributed directly and indirectly to this research. Declaration of Conflicting Interests: No potential competing interest was reported by the authors. Funding: This work was supported by the NIH under Grants K01 MD014177, R24 AG063724, and P30 AG072973.

## Abbreviations

(ADRD): Alzheimer’s disease and related disorders
(CLAS): Culturally and linguistically appropriate services
(GPACS-D): General Practitioners Confidence and Attitude Scale for Dementia
(KU ADRC): University of Kansas Alzheimer’s Disease Research Center
(NAPA): National Alzheimer’s Project Act
(PCPs): Primary care providers

## References

Administration for Community Living. (2015). A statistical profile of older Hispanic Americans. https://www.acl.gov/sites/default/files/Aging%20and%20Disability%20in%20America/Statistical-Profile-Older-Hispanic-Ameri.pdf

Administration on Aging, Administration for Community Living, & US Department of Health & Human Services. (2016). A profile of older Americans: 2016.

Alzheimer’s Association. (2019). 2019 Alzheimer’s disease facts and figures: Special report: Alzheimer’s detection in the primary care setting: Connecting patients with physicians. Alzheimer’s & Dementia, 15(3), 321–387.

Alzheimer’s Association. (2021). 2021 Alzheimer’s disease facts and figures. Alzheimer’s & Dementia, 17(3).

Association of American Medical Colleges. (2017). Section III: Geographic distribution of the physician workforce by race and ethnicity (Diversity in the physician workforce: facts & figures 2014, Issue.

Barker, W. W., Luis, C., Harwood, D., Loewenstein, D., Bravo, M., Ownby, R., & Duara, R. (2005). The effect of a memory screening program on the early diagnosis of Alzheimer disease. Alzheimer Disease & Associated Disorders, 19(1), 1–7.

Bédard, M., Koivuranta, A., & Stuckey, A. (2004). Health impact on caregivers of providing informal care to a cognitively impaired older adult: rural versus urban settings. Canadian Journal of Rural Medicine, 9(1), 15–23.

Bentley, M. W., Kerr, R., Ginger, M., & Karagoz, J. (2019). Behavioural change in primary care professionals undertaking online education in dementia care in general practice. Australian journal of primary health, 25(3), 244–249.

Black, C. M., Fillit, H., Xie, L., Hu, X., Kariburyo, M. F., Ambegaonkar, B. M., Baser, O., Yuce, H., & Khandker, R. K. (2018). Economic burden, mortality, and institutionalization in patients newly diagnosed with Alzheimer’s disease. Journal of Alzheimer’s Disease, 61(1), 185–193.

Boise, L., Camicioli, R., Morgan, D. L., Rose, J. H., & Congleton, L. (1999). Diagnosing dementia: perspectives of primary care physicians. The Gerontologist, 39(4), 457–464.

Bradford, A., Kunik, M. E., Schulz, P., Williams, S. P., & Singh, H. (2009). Missed and delayed diagnosis of dementia in primary care: prevalence and contributing factors. Alzheimer disease and associated disorders, 23(4), 306.

Cherry, D. L., Vickrey, B. G., Schwankovsky, L., Heck, E., Plauchm, M., & Yep, R. (2004). Interventions to improve quality of care: the Kaiser Permanente-alzheimer’s Association Dementia Care Project. Am J Manag Care, 10(8), 553–560.

Chodosh, J., Berry, E., Lee, M., Connor, K., DeMonte, R., Ganiats, T., Heikoff, L., Rubenstein, L., Mittman, B., & Vickrey, B. (2006). Effect of a dementia care management intervention on primary care provider knowledge, attitudes, and perceptions of quality of care. Journal of the American Geriatrics Society, 54(2), 311–317.

Chodosh, J., Mittman, B. S., Connor, K. I., Vassar, S. D., Lee, M. L., DeMonte, R. W., Ganiats, T. G., Heikoff, L. E., Rubenstein, L. Z., & Della Penna, R. D. (2007). Caring for patients with dementia: how good is the quality of care? Results from three health systems. Journal of the American Geriatrics Society, 55(8), 1260–1268.

Cooper-Patrick, L., Gallo, J. J., Gonzales, J. J., Vu, H. T., Powe, N. R., Nelson, C., & Ford, D. E. (1999). Race, gender, and partnership in the patient-physician relationship. Jama, 282(6), 583–589. https://doi.org/10.1001/jama.282.6.583

Cooper, L. A., Roter, D. L., Johnson, R. L., Ford, D. E., Steinwachs, D. M., & Powe, N. R. (2003). Patient-centered communication, ratings of care, and concordance of patient and physician race. Ann Intern Med, 139(11), 907–915. https://doi.org/10.7326/0003-4819-139-11-200312020-00009

Costa, G. D. d., Spineli, V. M. C. D., & Oliveira, M. A. d. C. (2019). Professional education on dementias in Primary Health Care: an integrative review. Revista brasileira de enfermagem, 72, 1086–1093.

Cotter, V. T. (2006). Alzheimer’s disease: issues and challenges in primary care. Nursing Clinics, 41(1), 83–93.

Drabo, E. F., Barthold, D., Joyce, G., Ferido, P., Chui, H. C., & Zissimopoulos, J. (2019). Longitudinal analysis of dementia diagnosis and specialty care among racially diverse Medicare beneficiaries. Alzheimer’s & Dementia, 15(11), 1402–1411.

Edwards, R., Voss, S. E., & Iliffe, S. (2015). The development and evaluation of an educational intervention for primary care promoting person-centred responses to dementia. Dementia, 14(4), 468–482.

Ferraro, F. R. (2015). Minority and cross-cultural aspects of neuropsychological assessment: enduring and emerging trends. Psychology Press.

Galvin, J. E., Meuser, T. M., & Morris, J. C. (2012). Improving physician awareness of Alzheimer’s disease and enhancing recruitment: the Clinician Partners Program. Alzheimer disease and associated disorders, 26(1), 61.

Galvin, J. E., Tolea, M. I., & Chrisphonte, S. (2020). What older adults do with the results of dementia screening programs. PLoS One, 15(7), e0235534.

Ganguli, I., Souza, J., McWilliams, J. M., & Mehrotra, A. (2018). Practices caring for the underserved are less likely to adopt Medicare’s annual wellness visit. Health affairs, 37(2), 283–291.

Gitlin, L. N., Kales, H. C., & Lyketsos, C. G. (2012). Nonpharmacologic management of behavioral symptoms in dementia. Jama, 308(19), 2020–2029.

Harris, P. A., Taylor, R., Thielke, R., Payne, J., Gonzalez, N., & Conde, J. G. (2009). Research electronic data capture (REDCap)—a metadata-driven methodology and workflow process for providing translational research informatics support. Journal of biomedical informatics, 42(2), 377–381.

Hebert, L. E., Weuve, J., Scherr, P. A., & Evans, D. A. (2013). Alzheimer disease in the United States (2010–2050) estimated using the 2010 census. Neurology, 80(19), 1778–1783.

Hurd, M. D., Martorell, P., Delavande, A., Mullen, K. J., & Langa, K. M. (2013). Monetary costs of dementia in the United States. New England Journal of Medicine, 368(14), 1326–1334.

Institute of Medicine. (2001). National Academies Press (US)

Copyright 2001 by the National Academy of Sciences. All rights reserved. https://doi.org/10.17226/10027

King, W. D., Wong, M. D., Shapiro, M. F., Landon, B. E., & Cunningham, W. E. (2004). Does racial concordance between HIV-positive patients and their physicians affect the time to receipt of protease inhibitors? J Gen Intern Med, 19(11), 1146–1153. https://doi.org/10.1111/j.1525-1497.2004.30443.x

Koller, D., Hua, T., & Bynum, J. P. (2016). Treatment Patterns with Antidementia Drugs in the United States: Medicare Cohort Study. J Am Geriatr Soc, 64(8), 1540–1548. https://doi.org/10.1111/jgs.14226

Lathren, C. R., Sloane, P. D., Hoyle, J. D., Zimmerman, S., & Kaufer, D. I. (2013). Improving dementia diagnosis and management in primary care: a cohort study of the impact of a training and support program on physician competency, practice patterns, and community linkages. BMC geriatrics, 13(1), 1–7.

Lee, L., Weston, W. W., & Hillier, L. M. (2013). Developing memory clinics in primary care: an evidence-based interprofessional program of continuing professional development. Journal of Continuing Education in the Health Professions, 33(1), 24–32.

Li, H. (2012). Unmet service needs: a comparison between dementia and non-dementia caregivers. Home health care services quarterly, 31(1), 41–59.

Lin, P. J., Emerson, J., Faul, J. D., Cohen, J. T., Neumann, P. J., Fillit, H. M., Daly, A. T., Margaretos, N., & Freund, K. M. (2020). Racial and Ethnic Differences in Knowledge About One’s Dementia Status. Journal of the American Geriatrics Society, 68(8), 1763–1770.

Martinez, K. M., Green, C. E., & Sanudo, F. M. (2004). The CLAS challenge: promoting culturally and linguistically appropriate services in health care. International Journal of Public Administration, 27(1-2), 39–61.

Mason, R., Doherty, K., Eccleston, C., Annear, M., Lo, A., Tierney, L., McInerney, F., & Robinson, A. (2019). General practitioners attitude and confidence scale for dementia (GPACS-D): confirmatory factor analysis and comparative subscale scores among GPs and supervisors. BMC family practice, 20(1), 1–8.

Mason, R., Doherty, K., Eccleston, C., Winbolt, M., Long, M., & Robinson, A. (2020). Effect of a dementia education intervention on the confidence and attitudes of general practitioners in Australia: a pretest post-test study. BMJ open, 10(1), e033218.

Mesterton, J., Wimo, A., Langworth, S., Winblad, B., & Jonsson, L. (2010). Cross sectional observational study on the societal costs of Alzheimer’s disease. Current Alzheimer Research, 7(4), 358–367.

Milne, A., Culverwell, A., Guss, R., Tuppen, J., & Whelton, R. (2008). Screening for dementia in primary care: a review of the use, efficacy and quality of measures. International psychogeriatrics, 20(5), 911–926.

Ortiz, F., & Fitten, L. J. (2000). Barriers to healthcare access for cognitively impaired older Hispanics. Alzheimer Disease & Associated Disorders, 14(3), 141–150.

Pan American Health Organization. (2021). Leading causes of mortality and health loss at the regional, subregional, and country levels in the Region of the Americas, 2000-2019.

Perry, M., Drašković, I., Lucassen, P., Vernooij-Dassen, M., van Achterberg, T., & Rikkert, M. O. (2011). Effects of educational interventions on primary dementia care: a systematic review. International journal of geriatric psychiatry, 26(1), 1–11.

Persky, S., Kaphingst, K. A., Allen, V. C., Jr., & Senay, I. (2013). Effects of patient-provider race concordance and smoking status on lung cancer risk perception accuracy among African-Americans. Ann Behav Med, 45(3), 308–317. https://doi.org/10.1007/s12160-013-9475-9

Polese, F., Moretta Tartaglione, A., & Cavacece, Y. (2016). Patient empowerment for healthcare service quality improvements: a value co-creation view. proceedings 19th Toulon-Verona International Conference Excellence in Services,

Rosen, C. S., Chow, H. C., Greenbaum, M. A., Finney, J. F., Moos, R. H., Sheikh, J. I., & Yesavage, J. A. (2002). How well are clinicians following dementia practice guidelines? Alzheimer Disease & Associated Disorders, 16(1), 15–23.

Rosenstock, I. M., Strecher, V. J., & Becker, M. H. (1988). Social learning theory and the health belief model. Health education quarterly, 15(2), 175–183.

Saha, S., & Beach, M. C. (2020). Impact of physician race on patient decision-making and ratings of physicians: a randomized experiment using video vignettes. Journal of general internal medicine, 1-8.

Saha, S., Komaromy, M., Koepsell, T. D., & Bindman, A. B. (1999). Patient-physician racial concordance and the perceived quality and use of health care. Archives of Internal Medicine, 159(9), 997–1004.

Sallim, A. B., Sayampanathan, A. A., Cuttilan, A., & Ho, R. C.-M. (2015). Prevalence of mental health disorders among caregivers of patients with Alzheimer disease. Journal of the American Medical Directors Association, 16(12), 1034–1041.

Scharlach, A. E., Giunta, N., Chow, J. C.-C., & Lehning, A. (2008). Racial and ethnic variations in caregiver service use. Journal of aging and health, 20(3), 326–346.

Traylor, A. H., Schmittdiel, J. A., Uratsu, C. S., Mangione, C. M., & Subramanian, U. (2010). Adherence to cardiovascular disease medications: does patient-provider race/ethnicity and language concordance matter? J Gen Intern Med, 25(11), 1172–1177. https://doi.org/10.1007/s11606-010-1424-8

U.S. Department of Health & Human Services. (2016). National Plan to Address Alzheimer’s Disease: 2016 Update. Office of The Assistant Secretary for Planning and Evaluation. https://aspe.hhs.gov/national-plan-address-alzheimers-disease-2015-update#goal4

U.S. Department of Health and Human Services. (2017). Summary health statistics tables for the U.S. Population: National Health Interview Survey, 2016. https://ftp.cdc.gov/pub/Health_Statistics/NCHS/NHIS/SHS/2016_SHS_Table_P-11.pdf

US Department of Health & Human Services. (2018). A physician’s practical guide to culturally competent care. Retrieved 8/28/2018 from https://www.thinkculturalhealth.hhs.gov/education/physicians

Vega, W. A., Kolody, B., Aguilar-Gaxiola, S., & Catalano, R. (1999). Gaps in service utilization by Mexican Americans with mental health problems. American Journal of Psychiatry, 156(6), 928–934.

Vollmar, H. C., Grässel, E., Lauterberg, J., Neubauer, S., Grossfeld-Schmitz, M., Koneczny, N., Schürer-Maly, C.-C., Koch, M., Ehlert, N., & Holle, R. (2007). Multimodal training of general practitioners--evaluation and knowledge increase within the framework of the dementia management initiative in general medicine (IDA). Zeitschrift fur arztliche Fortbildung und Qualitatssicherung, 101(1), 27–34.

Wang, Y., Xiao, L. D., Ullah, S., He, G.-P., & De Bellis, A. (2017). Evaluation of a nurse-led dementia education and knowledge translation programme in primary care: a cluster randomized controlled trial. Nurse education today, 49, 1–7.

Weden, M. M., Shih, R. A., Kabeto, M. U., & Langa, K. M. (2018). Secular Trends in Dementia and Cognitive Impairment of US Rural and Urban Older Adults. American journal of preventive medicine, 54(2), 164–172.

Wimo, A. (2004). Cost effectiveness of Cholinesterase Inhibitors in the treatment of Alzheimer’s Disease. Drugs & aging, 21(5), 279–295.

Winblad, B., & Wimo, A. (1999). Assessing the societal impact of acetylcholinesterase inhibitor therapies. Alzheimer disease and associated disorders, 13, S9–19.

